# “Clinical Significance and Distribution of Cerebrospinal Fluid Type IV Oligoclonal Band Pattern in Neurological Disorders: A Retrospective Analysis”

**DOI:** 10.64898/2026.10.27.26365939

**Authors:** Lucie Lefeuvre, Nathalie Lambeng, Christine Deffert, Patrice H. Lalive, Gautier Breville

**Affiliations:** Department of Clinical Neurosciences, Division of Neurology, Unit of Neuroimmunology, University Hospitals of Geneva, Geneva, Switzerland; Department of Laboratory Medicine, University Hospitals of Geneva, Geneva, Switzerland; Department of Pathology and Immunology, Faculty of Medicine, University of Geneva, Geneva, Switzerland

**Keywords:** type IV oligoclonal band pattern, mirror oligoclonal band pattern

## Abstract

**Background and Objectives:** To characterize the clinical features and cerebrospinal fluid (CSF) findings of patients exhibiting a type IV oligoclonal band (OCB) pattern, defined by the presence of identical IgG bands in both CSF and serum.

**Methods:** This retrospective observational study included all consecutive adult patients with a type IV pattern detected during routine CSF analysis at Geneva University Hospitals in 2023. Demographic, clinical, and laboratory data were collected through chart review.

**Results:** A total of 178 patients were included. The mean age was 60.2 years (SD 16.5), and 51.7% were female. The type IV pattern was most frequently observed in non-inflammatory neurological diseases (NIND) (n=103, 57.9%). Other inflammatory neurological diseases accounted for 37 cases (20.7%), multiple sclerosis was identified in 8 patients (4.5%), and peripheral nervous system disorders were present in 20 patients (11.2%). Systemic inflammatory comorbidities were observed in 40 patients (22.5%). An elevated albumin quotient was present in 73 patients (41%) and was most common among those with CNS infections, aseptic meningitis, CNS neoplastic diseases, and peripheral nervous system disorder. Repeated CSF analyses demonstrated temporal fluctuations in the type IV pattern in some patients, without corresponding changes in diagnosis.

**Discussion:** The type IV pattern is predominantly associated with NIND and is relatively uncommon in multiple sclerosis. Although it may reflect systemic immune activation and/or blood–brain barrier dysfunction in certain inflammatory conditions, its clinical significance remains nonspecific. These findings support cautious interpretation of the type IV pattern in routine clinical practice.

## Introduction

The presence of cerebrospinal fluid (CSF)-restricted oligoclonal band (OCB) reflects intrathecal IgG synthesis and is widely recognized as an important biomarker of central nervous system (CNS) inflammation. Detection of intrathecal OCB is primarily used to support the diagnosis of multiple sclerosis (MS) according to the 2024 McDonald criteria^1^. Similarly, CSF-restricted OCB can be detected in several other neurological conditions, including autoimmune encephalitis, systemic autoimmune diseases with CNS involvement, and CNS infections^2^.

In contrast, the non-CSF restricted OCB–defined as identical bands present in both CSF and serum, also referred to as the type IV pattern–have uncertain diagnostic significance. This pattern is thought to reflect the entry of a systemic humoral immune response into the CNS, across the blood-brain barrier (BBB)^3^. A limited number of case reports and small studies have described the type IV pattern in a range of conditions. In inflammatory neurological diseases, such as autoimmune encephalitis, acute disseminated encephalomyelitis (ADEM), myelin oligodendrocyte glycoprotein antibody–associated disease (MOGAD), and neuromyelitis optica spectrum disorder (NMOSD), this pattern has been reported, suggesting that the pathogenic autoantibodies involved are primarily produced in the periphery^3^. The type IV pattern has also been described in auto-immune systemic diseases, including systemic lupus erythematosus (SLE), sarcoidosis and Sjögren syndrome, even in the absence of associated neurological involvement^4^. Morevover, the type IV pattern is well documented in Guillain–Barré syndrome (GBS) and chronic inflammatory demyelinating polyneuropathy (CIDP) where it reflects BBB disruption^5^. Finally, for reasons that remain unclear, this pattern is not uncommon in non-inflammatory neurological conditions such as neurodegenerative or vascular diseases^4,6^.

Because OCB analysis is widely used in the diagnostic work-up of suspected CNS inflammatory disease, the type IV pattern has been observed across a broad range of conditions. However, its clinical significance remains unclear due to the limited availability of well-characterized clinical data. The aim of this study was to describe the clinical characteristics and CSF findings associated with type IV pattern and to assess its clinical utility.

## Methods

### Patients

This retrospective observational study included all consecutive adult patients (≥18 years) with a type IV pattern detected on CSF analysis performed as part of routine care at Geneva University Hospitals, between 1 January 2023 and 31 December 2023. The study protocol was approved by the Swiss Ethics Committee (No. 2020-02822).

All eligible patients underwent a chart review. Patients were excluded if clinical data were missing or if the CSF red blood cell count exceeded 1000/µL (risk to contaminate the results). Demographic data, clinical diagnoses retained one year after OCB analysis in CSF, and systemic comorbidities were collected. Each patient was assigned to a diagnostic category and, when appropriate, a subcategory (Table).

**Table.**
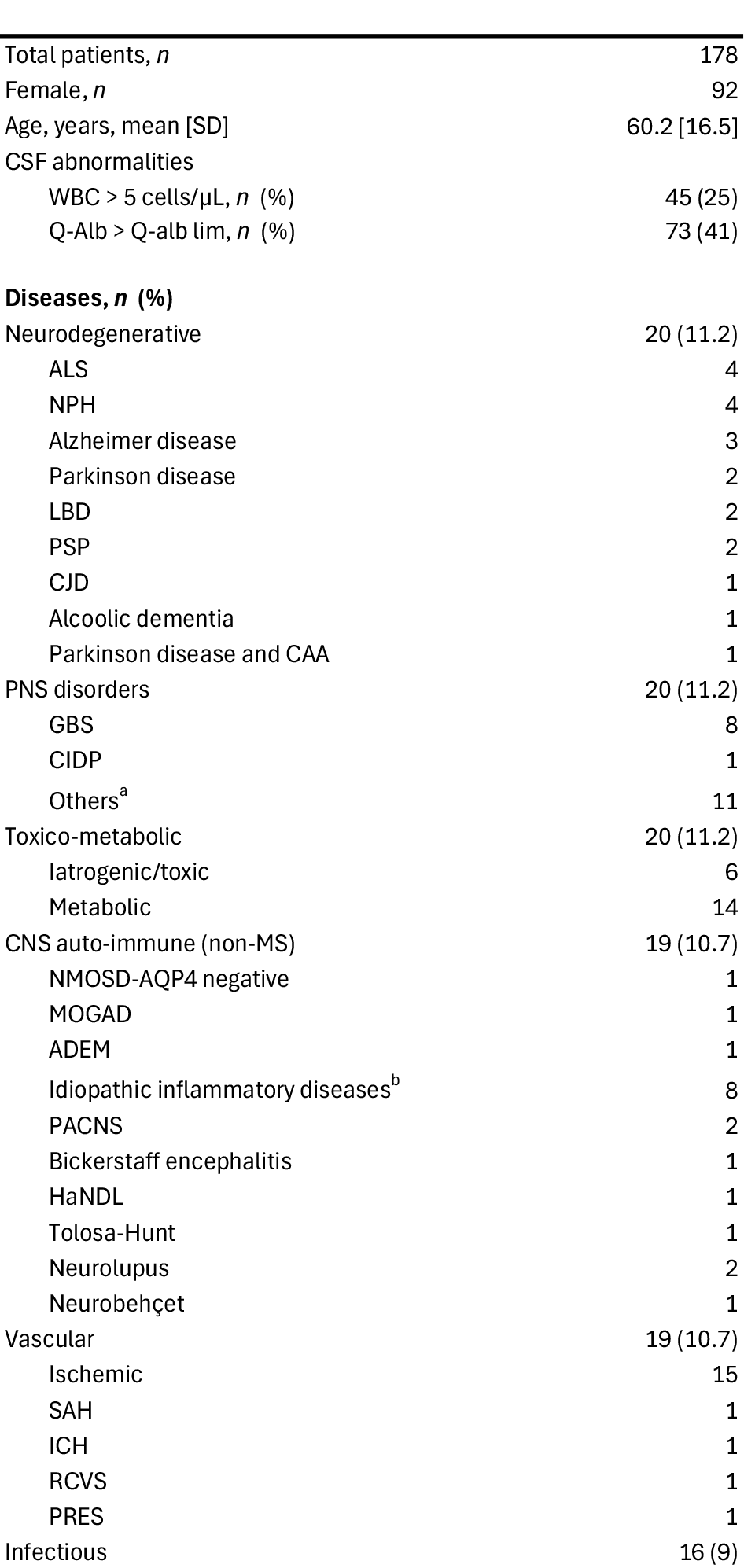
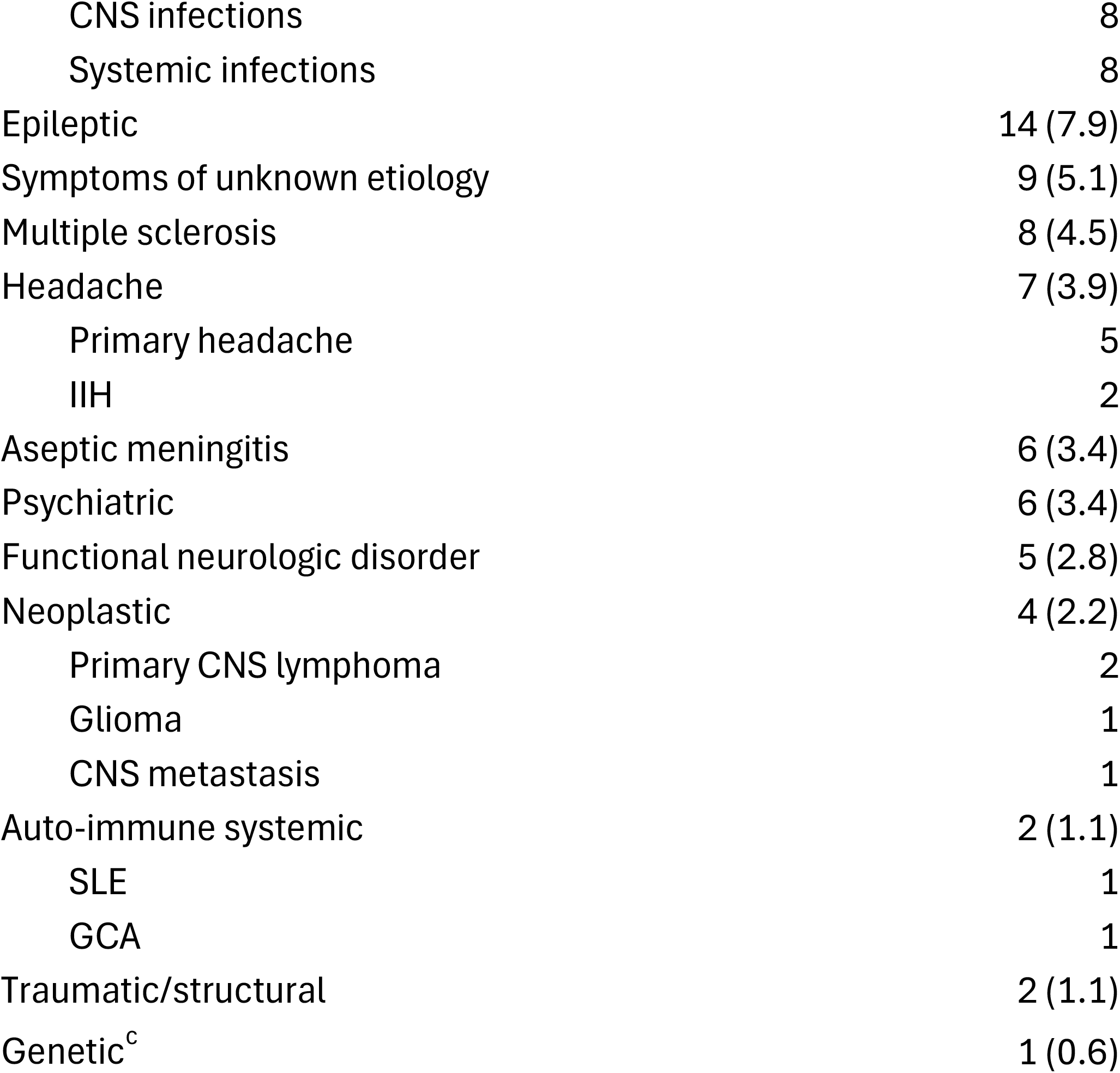
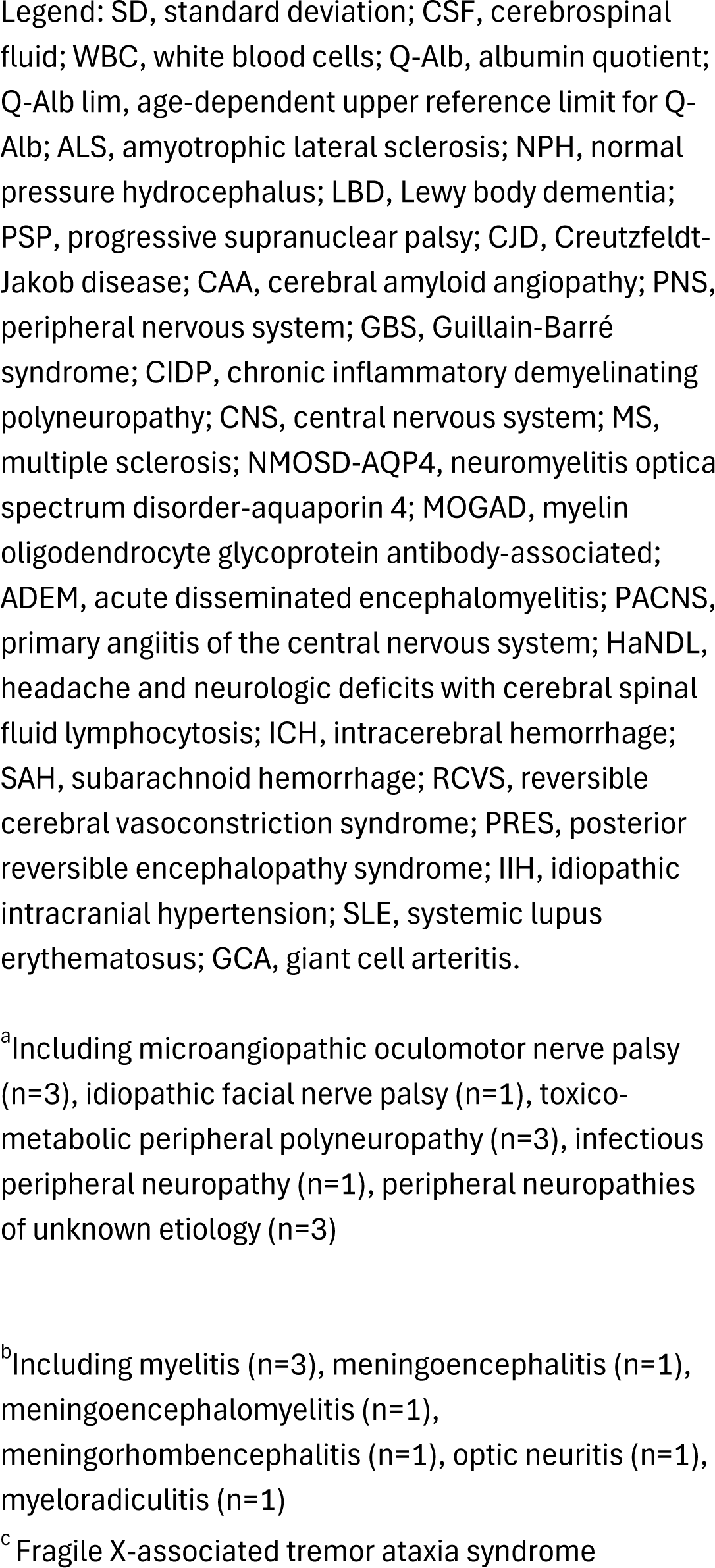
Demographics and clinical data.

**Table. Demographics and clinical data**
|  |  |
| --- | --- |
| Total patients, *n* | 178 |
| Female, *n* | 92 |
| Age, years, mean [SD] | 60.2 [16.5] |
| CSF abnormalities |  |
| WBC > 5 cells/μL, *n* (%) | 45 (25) |
| Q-Alb > Q-alb lim, *n* (%) | 73 (41) |
| **Diseases, *n* (%)** |  |
| Neurodegenerative | 20 (11.2) |
| ALS | 4 |
| NPH | 4 |
| Alzheimer disease | 3 |
| Parkinson disease | 2 |
| LBD | 2 |
| PSP | 2 |
| CJD | 1 |
| Alcoholic dementia | 1 |
| Parkinson disease and CAA | 1 |
| PNS disorders | 20 (11.2) |
| GBS | 8 |
| CIDP | 1 |
| Others<sup>a</sup> | 11 |
| Toxico-metabolic | 20 (11.2) |
| Iatrogenic/toxic | 6 |
| Metabolic | 14 |
| CNS auto-immune (non-MS) | 19 (10.7) |
| NMOSD-AQP4 negative | 1 |
| MOGAD | 1 |
| ADEM | 1 |
| Idiopathic inflammatory diseases<sup>b</sup> | 8 |
| PACNS | 2 |
| Bickerstaff encephalitis | 1 |
| HaNDL | 1 |
| Tolosa-Hunt | 1 |
| Neurolupus | 2 |
| Neurobehçet | 1 |
| Vascular | 19 (10.7) |
| Ischemic | 15 |
| SAH | 1 |
| ICH | 1 |
| RCVS | 1 |
| PRES | 1 |
| Infectious | 16 (9) |
| CNS infections | 8 |
| Systemic infections | 8 |
| Epileptic | 14 (7.9) |
| Symptoms of unknown etiology | 9 (5.1) |
| Multiple sclerosis | 8 (4.5) |
| Headache | 7 (3.9) |
| Primary headache | 5 |
| IIH | 2 |
| Aseptic meningitis | 6 (3.4) |
| Psychiatric | 6 (3.4) |
| Functional neurologic disorder | 5 (2.8) |
| Neoplastic | 4 (2.2) |
| Primary CNS lymphoma | 2 |
| Glioma | 1 |
| CNS metastasis | 1 |
| Auto-immune systemic | 2 (1.1) |
| SLE | 1 |
| GCA | 1 |
| Traumatic/structural | 2 (1.1) |
| Genetic<sup>c</sup> | 1 (0.6) |

Legend: SD, standard deviation; CSF, cerebrospinal fluid; WBC, white blood cells; Q-Alb, albumin quotient; Q-Alb lim, age-dependent upper reference limit for Q-Alb; ALS, amyotrophic lateral sclerosis; NPH, normal pressure hydrocephalus; LBD, Lewy body dementia; PSP, progressive supranuclear palsy; CJD, Creutzfeldt-Jakob disease; CAA, cerebral amyloid angiopathy; PNS, peripheral nervous system; GBS, Guillain-Barré syndrome; CIDP, chronic inflammatory demyelinating polyneuropathy; CNS, central nervous system; MS, multiple sclerosis; NMOSD-AQP4, neuromyelitis optica spectrum disorder-aquaporin 4; MOGAD, myelin oligodendrocyte glycoprotein antibody-associated; ADEM, acute disseminated encephalomyelitis; PACNS, primary angiitis of the central nervous system; HaNDL, headache and neurologic deficits with cerebral spinal fluid lymphocytosis; ICH, intracerebral hemorrhage; SAH, subarachnoid hemorrhage; RCVS, reversible cerebral vasoconstriction syndrome; PRES, posterior reversible encephalopathy syndrome; IIH, idiopathic intracranial hypertension; SLE, systemic lupus erythematosus; GCA, giant cell arteritis.
<sup>a</sup>Including microangiopathic oculomotor nerve palsy (n=3), idiopathic facial nerve palsy (n=1), toxicometabolic peripheral polyneuropathy (n=3), infectious peripheral neuropathy (n=1), peripheral neuropathies of unknown etiology (n=3)
<sup>b</sup>Including myelitis (n=3), meningoencephalitis (n=1), meningoencephalomyelitis (n=1), meningorhombencephalitis (n=1), optic neuritis (n=1), myeloradiculitis (n=1)
<sup>c</sup>Fragile X-associated tremor ataxia syndrome

### CSF Analysis

CSF white blood cell (WBC) counts were recorded and considered abnormal if >5 cells/µL. BBB integrity was assessed using the albumin quotient (Q-Alb), calculated as the ratio of CSF albumin to serum albumin. An elevated Q-Alb was defined as a value above the age-dependent upper reference limit, calculated according to the formula (age in years/15)+4^7^.

All OCB analyses were performed in the Geneva University Hospitals laboratory using isoelectric focusing combined with IgG immunofixation (kit Hydrasis Sebia, Evry, France) on a semi-automated platform. Paired serum and CSF samples were run side by side on an agarose gel. Immunofixation with enzyme-labeled anti-IgG was used to detect IgG clones. Results were interpreted by two experienced laboratory biologists who were blinded to the clinical diagnoses. In equivocal cases, a consensus was reached between the two biologists. The type IV pattern corresponds to identical OCB in both CSF and serum, and contrasts with the other patterns: type I (no OCB), type II (at least two CSF-restricted OCB), type III (identical OCB in CSF and serum with at least two additional CSF-restricted OCB), and type V (monoclonal bands identical in both CSF and serum)^5^.

If a patient underwent multiple lumbar punctures (LPs) during the study period, the results from the first LP were included in the primary analysis (Table). Evolution of the OCB pattern observed in subsequent LPs was next described.

## Results

### Patient characteristics

Between January 1^st^ 2023 and December 31^th^ 2023, 207 adult patients with a type IV OCB pattern were identified at Geneva University Hospitals. Twenty-one patients were excluded due to missing clinical data, and an additional eight were excluded because of CSF red blood cell counts exceeding 1000/µL, leaving 178 patients in the final analysis (Figure 1 - Flowchart). Table and Figure 2 summarize the demographic characteristics, diagnostic categories and subcategories, and CSF findings. Of the included patients, 51.7% were female, and the mean age was 60.2 years (SD 16.5).

**Figure 1.**
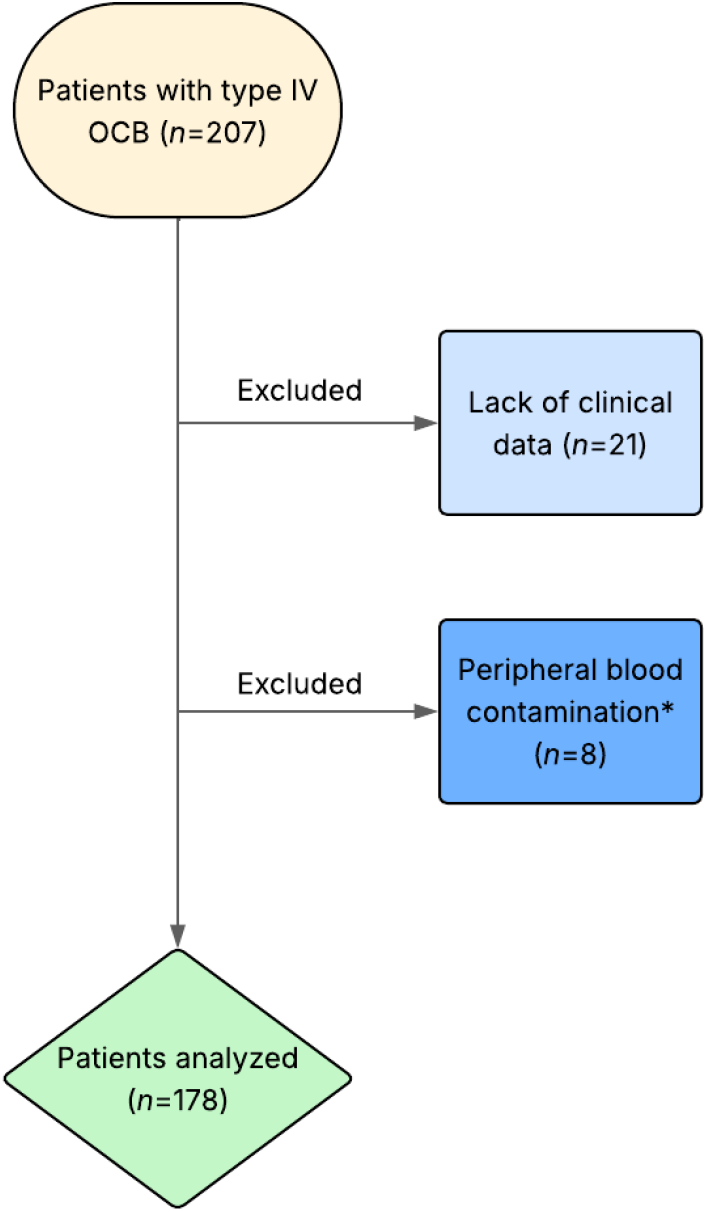
Flowchart of study patient selection Legend: *Cerebrospinal fluid red blood cells > 1000/µL

**Figure 2.**
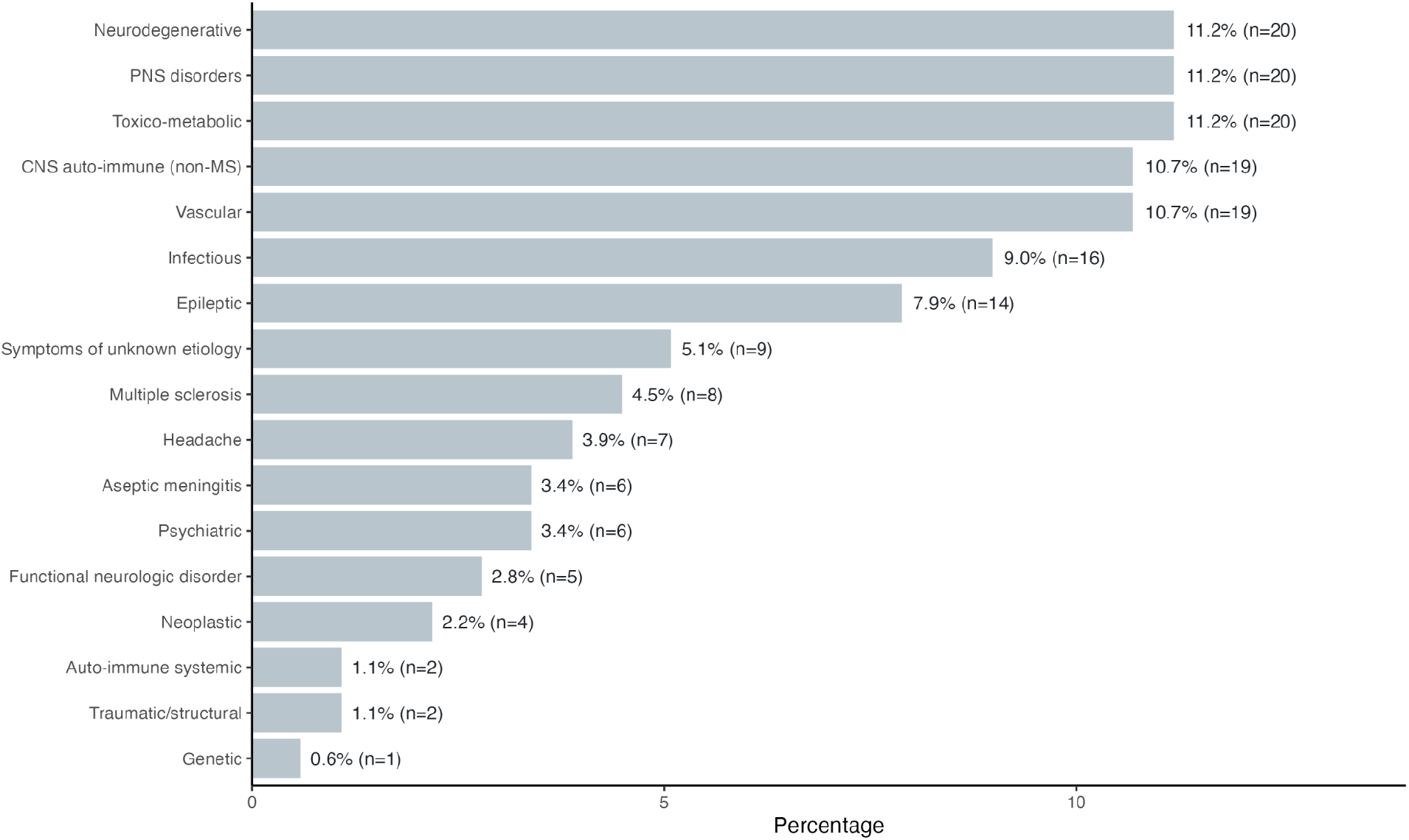
Etiologies of type IV OCB pattern Legend: OCB, oligoclonal bands; PNS, Peripheral nervous system; CNS, Central nervous system; MS, Multiple sclerosis.

### Clinical diagnoses associated with type IV pattern

The type IV pattern was most commonly observed in non-inflammatory neurological diseases (NIND) (n = 103, 57.9%), including neurodegenerative (11.2%), toxicometabolic (11.2%), vascular (10.7%), epileptic (7.9%), psychiatric (3.4%), functional (3.4%), unknown origin (5.1%), headache-related (3.9%), traumatic or structural (1.1%), and genetic (0.6%) disorders. The type IV pattern was also observed in patients with CNS inflammatory diseases including, on the one hand, patients with MS diagnosed according to the 2024 McDonald criteria (n = 8, 4.5%), and, on the other hand, patients with other inflammatory neurological diseases (OIND) (n = 37, 20.7%). The latter group comprised 19 CNS autoimmune diseases, 8 CNS infections, 6 cases of aseptic meningitis, and 4 CNS neoplastic diseases. Finally, the type IV pattern was identified in peripheral nervous system (PNS) disorders (n = 20, 11.2%), including 8 cases of GBS and 1 case of CIDP (Table and Figure 2).

### Systemic comorbidities

A total of 40 patients (22.5%) had systemic inflammatory comorbidity, including systemic autoimmune diseases (n=16), autoimmune endocrinopathies (n=3), systemic neoplastic diseases (n=13), and systemic infections (n=8) (Figure 3). Among these patients, some had neurological conditions unrelated to their comorbidity, while the others had no neurological disease.

**Figure 3.**
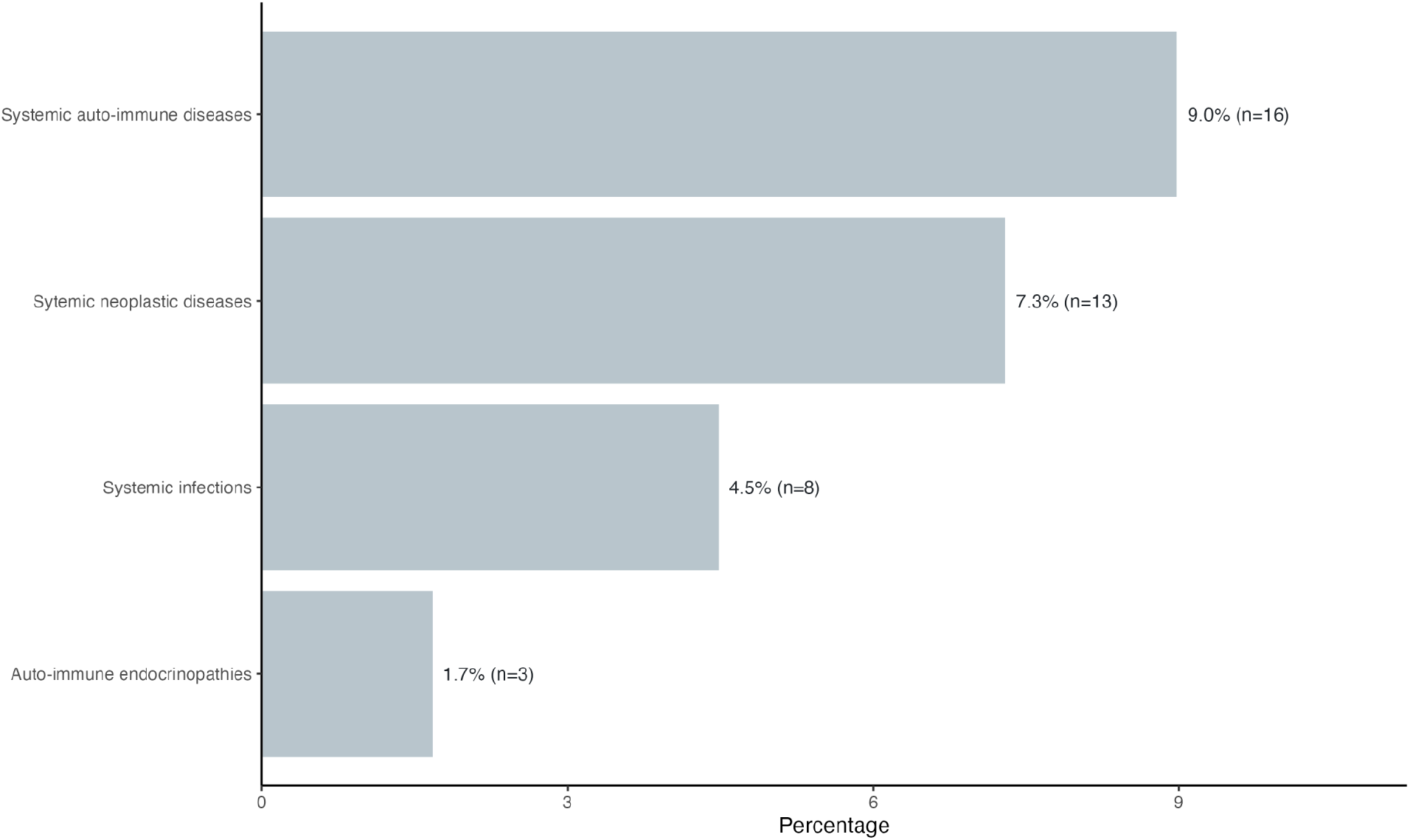
Association of systemic comorbidities with type IV CSF OCB pattern Legend: CSF, cerebrospinal fluid; OCB, oligoclonal bands.

### OCB pattern evolution over time

During the study period, 20 patients (11.2%) underwent repeated CSF examinations (i.e., two or more LPs). Changes in the OCB pattern over time were assessed. In 12 patients, the type IV pattern remained stable (mean interval between the first and last LP: 32.1 days, range 1–181 days). In 5 patients, the type IV pattern shifted to type I (mean interval: 16.4 days, range 6–31 days). In 3 patients—one with an autoimmune CNS disease, one with aseptic meningitis, and one with a CNS neoplastic disease— the type IV pattern shifted to type II or III (mean interval: 47.1 days, range 7–91 days). No changes in clinical diagnosis were observed between the first and last LP.

### CSF abnormalities

Overall, Q-Alb was elevated in 73 of 178 patients (41%) (Table). Elevated Q-Alb was most frequently observed in PNS disorders (13/20)—including GBS (6/8) and CIDP (1/1)—CNS infections (6/8), aseptic meningitis (4/6), and CNS neoplastic diseases (4/4) (Supplementary table). CSF WBC counts were elevated in all CNS infections (8/8), all cases of aseptic meningitis (6/6), and most CNS neoplastic diseases (3/4). In contrast, WBC elevation was observed in only a minority of PNS disorders (4/20) (Supplementary table). In the combined diagnostic subgroup of 18 patients including those with CNS infections (n=8), aseptic meningitis (n=6) and neoplastic diseases (n=4), CSF WBC increase and Q-Alb elevation were significantly correlated (Spearman ρ = 0.75, p < 0.001, Supplementary figure).

## Discussion

Our study, aimed to determine the clinical associations of the type IV pattern. More than half of the patients (57.9%) had NIND, whereas 20.7% had OIND. Compared with OIND group, the type IV pattern was relatively uncommon in MS (4.5%). These findings are consistent with those of a previous study including patients with a type IV pattern^3^. In that cohort, 59.9% had NIND, 32.3% had OIND, and 7.8% had MS. In our study, the type IV pattern was also observed in PNS disorders (11.2%), including GBS and CIDP. This is consistent with the existing literature, in which the type IV pattern has previously been reported in patients with PNS disorders, including GBS, CIDP, and other peripheral neuropathies^3,8^.

The type IV pattern is generally interpreted as reflecting a systemic humoral immune response, with passive transfer of serum IgG into the CSF through increased blood–brain barrier permeability^3,8^. In this context, OCB are thought to originate in the systemic circulation following repeated activation of the adaptive immune system, and subsequently enter the CNS due to increased BBB permeability^8^. Some authors have suggested that a history of systemic autoimmune disease or cancer may promote the presence of serum OCB^3^. In another cohort of patients with a type IV pattern, 37.8% had at least one autoimmune disease and 26.4% had a history of cancer^3^. In contrast, in our study, systemic autoimmune, oncological, or infectious comorbidities were present in less than one quarter of patients. This lower prevalence may reflect broader indications for lumbar puncture at our center and a bias of patients’ selection for LP should be considered. Our findings suggest that, in many patients, systemic OCB may result from past common infections, and do not necessarily indicate active systemic inflammatory diseases, as proposed by Pannewitz et al.^6^. According to this hypothesis, the reported increasing prevalence of the type IV pattern with age reflects the cumulative “immunological scars” of repeated infections over time^6^. The type IV pattern detection may therefore represent a long-term “fingerprint” of prior antigen exposure rather than current immune activity^6^.

Repeated CSF analyses were performed in 11.2% of patients with a range of neurological conditions. In 40% of them, the type IV pattern fluctuated over time. Fluctuations were not associated with subsequent changes in clinical diagnosis. Similar observations were reported in another cohort, in which 7.4% underwent repeated CSF analysis^3^. Because repeated LPs are relatively uncommon, data on this topic remain limited. The observed variability of the type IV pattern highlights the dynamic nature of systemic antibodies production. As these changes were not associated with clinical evolution, repeated CSF analysis appears to offer limited practical benefit^3^.

As the type IV pattern reflects the presence of serum OCB in the CSF, Q-Alb, a marker of BBB disruption, was evaluated. Elevated Q-Alb was associated with increased CSF WBC counts in highly inflammatory CNS conditions, including CNS infections, aseptic meningitis, and neoplastic diseases. Previous studies have shown that in inflammatory CNS disorders such as meningitis, BBB disruption is reflected by Q-Alb elevation, which correlates with pleocytosis^9^. Elevated Q-Alb was also observed in PNS disorders such as GBS and CIDP, where it is a well-established feature and part of the diagnostic criteria^5^. In these situations, it reflects BBB damage and is not associated with pleocytosis. However, most of all patients in our cohort did not have elevated Q-Alb despite the presence of the type IV pattern. This may be explained by the limited sensitivity of Q-Alb for detecting subtle BBB dysfunction, particularly when smaller serum molecules can cross the barrier^10^. In addition, aging is known to affect BBB integrity. The gradual decline in BBB function with age may contribute to increased passive diffusion of systemic antibodies into the CSF^6,11^.

Our study aimed to determine the clinical associations and diagnostic implications of type IV pattern. The single-center retrospective design of our study may introduce biases. However, its strength lies in the broad inclusion criteria, which provide a comprehensive overview of the clinical contexts in which the type IV pattern is observed. Further validation in larger, multicenter cohorts is needed to better define the clinical significance and utility of this laboratory finding.

## Data Availability

All data produced in the present study are available upon reasonable request to the authors

## Declaration of Generative AI and AI-assisted technologies in the writing process

Editing purposes

## Ethics approval

The study was approved by the local Institutional Review Board, and all participants provided written informed consent prior to inclusion according to the Declaration of Helsinki.

## Funding sources

None

## Declaration of competing interest

The authors declare that they have no conflicts of interest in relation to this work.

## Notes

### Competing Interest Statement

The authors have declared no competing interest.

### Author Declarations

Ethical approval was given by the Swiss Ethics Committee of Geneva (No. 2020-02822).

