## supplementary figure for "“Clinical Significance and Distribution of Cerebrospinal Fluid Type IV Oligoclonal Band Pattern in Neurological Disorders: A Retrospective Analysis”"

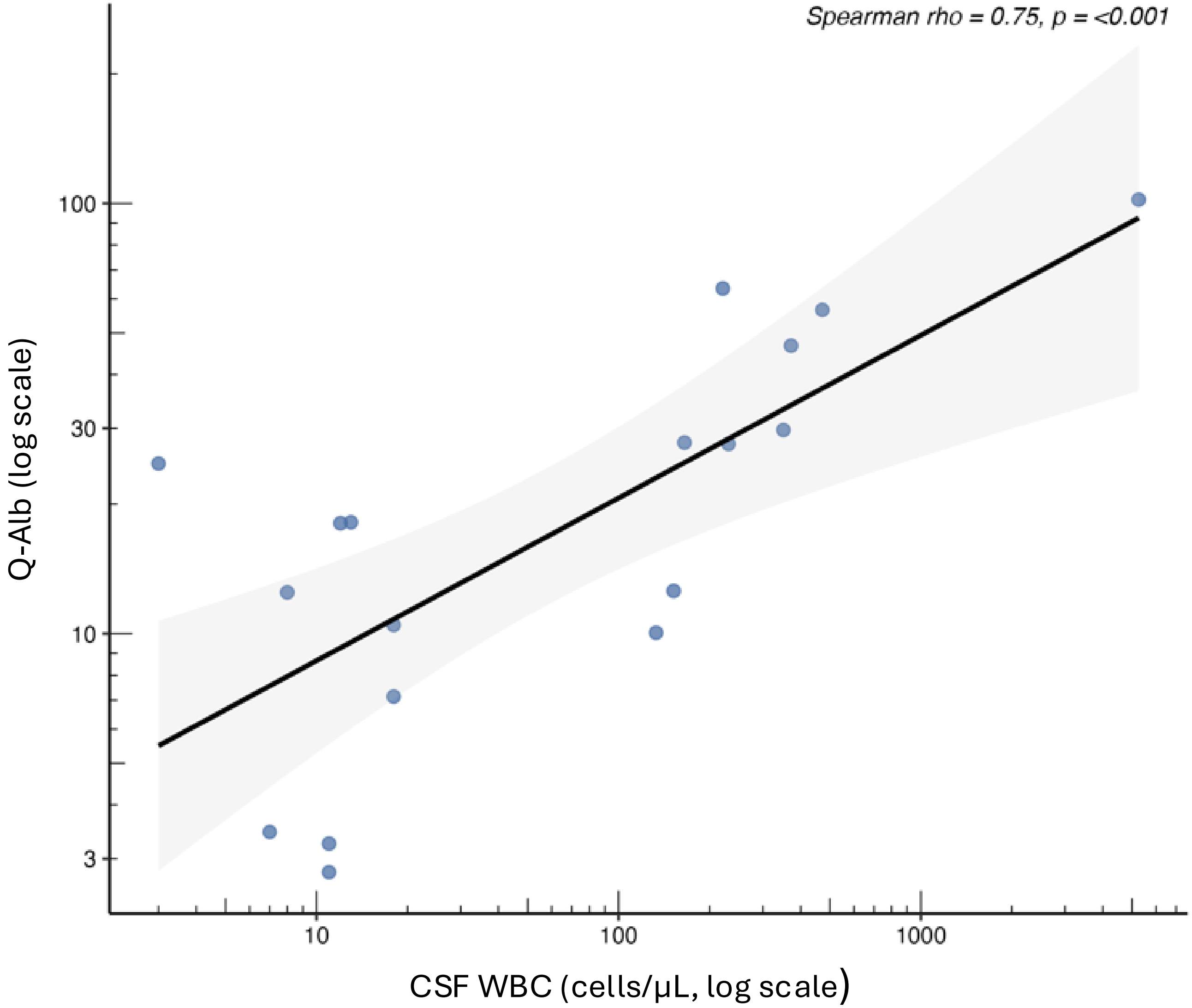


Supplementary figure. Correlation between Q-Alb elevation and CSF WBC increase in the CNS infections, aseptic meningitis and neoplastic groups.

Legend: Q-Alb, albumin quotient; CSF, cerebrospinal fluid; WBC, white blood cells; CNS, central nervous system.
