## supplementary table for "“Clinical Significance and Distribution of Cerebrospinal Fluid Type IV Oligoclonal Band Pattern in Neurological Disorders: A Retrospective Analysis”"

**Supplementary table. WBC and albumin quotient levels according to diseases categorie**

| **Diseases** | ***n* patients** | **WBC > 5 cells/µL, *n*** | **Q-Alb > Q-Alb lim, *n*** |
|---|---|---|---|
| Neurodegenerative | 20 | 0 | 4 |
| PNS disorders | 20 | 4 | 13 |
| Toxico-metabolic | 20 | 2 | 9 |
| CNS auto-immune (non-MS) | 19 | 9 | 9 |
| Vascular | 19 | 5 | 10 |
| Infectious | 16 | 10 | 9 |
| CNS infections | 8 | 8 | 6 |
| Systemic infections | 8 | 2 | 3 |
| Epileptic | 14 | 2 | 5 |
| Symptoms of unknown etiology | 9 | 0 | 2 |
| Multiple sclerosis | 8 | 2 | 0 |
| Headache | 7 | 0 | 1 |
| Aseptic meningitis | 6 | 6 | 4 |
| Psychiatric | 6 | 0 | 1 |
| Functional neurologic disorder | 5 | 1 | 1 |
| Neoplastic | 4 | 3 | 4 |
| Auto-immune systemic | 2 | 0 | 0 |
| Traumatic/structural | 2 | 1 | 1 |
| Genetic[a] | 1 | 0 | 0 |

Legend: WBC, white blood cells; Q-Alb, albumin quotient; Q-Alb lim, age-dependent upper reference limit for Q-Alb; PNS, peripheral nervous system; CNS, central nervous system; MS, multiple sclerosis.

[a] Fragile X-associated tremor ataxia syndrome

S
